# Land use density, land use mix, and walking: insight from a simple theoretical model

**DOI:** 10.1101/2023.11.29.23299138

**Authors:** Yong Yang

## Abstract

**Background:** Land use density and land use mix are the two most studied characteristics of the built environment and generally, it is believed that higher land use density and higher level of land use mix are associated with more walking, especially for utilitarian purposes. However, although a large number of studies including population density and land use mix have been conducted over the last two decades, overall, the result is still not very clear.

**Methods:** I developed a simple computer model to simulate the influence of land use density and land use mix on walking. It includes a hypothetical city with a grid space and a number of households and non-residential places located across the city. Eventually, we varied the contexts to explore the corresponding results of the walking trips and walking distance across the population.

**Results:** Only at a very low level, increased land use density may increase walking trips. At the same time, increased land use density may decrease the total distance of walking trips. A higher level of land use mix is associated with higher walking trips. However, a higher level of land use mix level may be not associated with a higher total distance of walking trips. The relationship between land use and walking is significantly modified by other factors.

**Conclusion:** The relationship between land use and walking is complicated because multiple factors at multiple levels could contribute to this relationship and the conclusions from the most existing practical studies may apply only to a specific population in a specific context. Computer models will be supplemental because they provide a method to disentangle the complex relationship by separating and removing confounding factors and focusing on the interested factors.

## 1. Introduction

Walking, as a subset of active travel, is influenced by various aspects of the built environment [1, 2]. Among these factors, land use density and land use mix stand out as the two extensively studied characteristics [3–9]. Both are considered part of the “3Ds”, which have been identified as significantly associated with walking and physical activity more broadly [9, 10]. Land use density involves quantifying the number of homes, people, or jobs within a given area. Density metrics encompass population density, employment density, and retail activity density. Land use density is straightforward to define and is convenient to measure. On the other hand, land use mix, or diversity, pertains to the variety of land uses present, encompassing residential, commercial, industrial, and other purposes. Unlike density, land use mix poses challenges in quantitative measurement. The two commonly used metrics for land use mix are entropy, reflecting the diversity of land uses in a neighborhood, and the dissimilarity index, which gauges the number of adjacent parcels with different uses [11, 12].

Generally, it is widely accepted that increased land use density [1, 13–15] and a higher level of land use mix [7, 15–17] are associated with elevated levels of walking (particularly for walking for utilitarian purposes), greater reliance on public transit, and a reduction in vehicle miles traveled (VMT). For example, as highlighted in the comprehensive review by Ewing and Cervero[13], doubling household or population density has been associated with a 7% increase in walking, while doubling the land use mix has shown a 15% increase in walking.

Residents of neighborhoods characterized by higher walkability—defined by factors such as increased residential density, land use mix, street connectivity, aesthetics, and safety—have been found to engage in an additional 70 minutes of physical activity and exhibit lower obesity prevalence compared to those in less walkable neighborhoods [18]. Notably, research indicates that the positive impact of higher land use density and mix extends beyond walking for transportation, also influencing recreational walking [17, 19, 20], as well as both work and non-work-related travel [21].

The relationships between land use density, land use mix, and walking may seem straightforward and intuitive. The rationale lies in the fact that higher density and land use mix contribute to increased proximity, effectively reducing distances between origins and destinations. Consequently, this reduction in trip distance may enhance the likelihood of individuals choosing walking as their preferred mode of travel. At the same time, higher density increases the visibility of others walking and fosters a sense of safety. Additionally, elevated density introduces traffic friction, leading to reduced traffic speeds, and higher land costs associated with increased density contribute to a decrease in parking supply and an increase in parking pricing, making driving less convenient [6]. A study [22] discovered that higher land use mix resulted in an overall increase in total trips, indicating induced travel. However, a significant portion of these additional trips were walking trips, leading to a decline in total vehicle travel. In contrast, lower density or reduced land use mix, characterized by the separation of uses into distinct residential, commercial, and industrial zones, may lead to increased travel distances, thereby discouraging walking trips.

Despite a substantial number of studies on population density and land use mix conducted over the last two decades, the overall results remain somewhat inconclusive. Several challenges and complexities contribute to the ambiguity in the findings. Firstly, conflicting conclusions have emerged from some studies. For instance, while density has been associated with specific purposes of walking, such as for travel and leisure, it may not exhibit a consistent relationship with overall walking or overall physical activity [5, 6]. Another example suggests that land use density and mix strongly influence walking for transportation but may not have the same impact on walking for recreation [23]. Secondly, even after accounting for numerous other factors, population and job densities have been found to exhibit weak associations with travel behavior [13]. This implies that land use density and mix might act as “proxies” for other, more influential factors. Thirdly, challenges related to measurement, especially concerning land use mix, contribute to the complexity. A review [24] indicated that land use mix had no effect on physical activity in 87% of the measures, with only 10% of measures showing an effect in the expected direction. Fourthly, the presence of a non-linear relationship between land use density and walking has been hypothesized and identified [3, 7]. For example, certain studies propose that density needs to surpass a specific threshold value to significantly alter travel mode share [25, 26]. Notably, in a national-level study in the United States [20], the positive relationship between population density and walking was only evident at extreme low and high density levels for recreational purposes. Conversely, for utilitarian purposes, the relationship was most pronounced at the highest density categories.

The relationships between land use density, land use mix, and walking are intricate and multifaceted. Firstly, density is often linked with other factors such as land use mix, the transit system, and parking management, collectively exerting impacts on travel patterns[27]. Increased density may enhance the cost efficiency of various facilities, including sidewalks, paths, and public transit [28]. This association is partly due to historical considerations, as many denser neighborhoods developed before 1950 were designed with a focus on multi-modal access, while numerous lower-density neighborhoods developed between 1950 and 2000 were designed with a car-oriented approach. Consequently, density serves as an indicator or proxy for other features [13]. For instance, higher density may be associated with characteristics such as lower-income residents, a lack of car ownership, and improved transit accessibility. Ultimately, the correlation between density and walking weakens when adjusted for covariates related to the built environment [13, 29]. Additionally, density and diversity are interconnected measures of the built environment that have been shown to influence physical activity [30]. Secondly, density is commonly quantified using metrics such as population density or employment density across various scales. It’s important to note that the relationship between land use density and mix can be influenced by the chosen scale of analysis [4, 31]. When data at a fine scale (e.g., parcel or census block level) are unavailable, researchers may resort to a broader scale (e.g., census tract or zipcode level), potentially limiting the ability to identify significant density variations within an area. Thirdly, the challenge of measurement is a significant factor, particularly in the case of land use mix [4, 11]. One notable issue is the absence of a standard definition and measurement approach for land use mix. This lack of standardization is evident in the lack of specificity in defining the categories of land uses considered and variations in the geographical scale used for measurement. The inconsistency in defining and measuring land use mix can introduce variability and make it challenging to compare findings across different studies. Fourthly, there is a trade-off between walking frequency and walking distance. Individuals in higher-density areas may engage in more walking trips, but these trips often tend to be shorter. For instance, research [31] suggests that high-density and mixed land use can contribute to a reduction in distances, subsequently lowering energy demands. This phenomenon arises because destinations are close to each other in areas characterized by high density and mixed land use. As a result, while individuals in these areas may walk more frequently, the shorter distances involved can impact the overall energy demands associated with walking.

The mixed results from previous empirical studies may be not surprising they are often specific to particular populations and contexts. However, it is important to identify a general pattern that could be applicable across various cases. To achieve this, a generic theoretical model can help to deepen our understanding of the general pattern, when complemented by existing empirical evidence. In Section 2 a computer model is described, and the model is developed to simulate how the pattern of walking varies by different scenarios of land use density and land use mix. Section 3 presents the simulated results, followed by Section 4, which discusses these results.

## 2. Methods

The model was developed using Java. It includes a hypothetical city with a size of 3.2 km × 3.2 km presented by a 320 × 320 grid space (i.e., each cell is 10m × 10m). A cell may be occupied by a household or a non-residential place, or be unoccupied. For simplifying purposes, we assume four categories of non-residential places (each category has an equal total number of non-residential places) serve people’s daily lives for various purposes. For example, these four categories may be interpreted as workplace, shop, restaurant, and social place. Trip length between a household to a certain non-residential place is measured by the Manhattan distance. We assume trip length is the only factor determining a person’s travel mode and walking or not obeys a simple dichotomous rule: if trip length is less than walking distance, walk; otherwise, not. Further, the same value of walking distance applies to every household in the model. For each scenario, we computed two measures. First, for each household, we added up the total number of categories if there was at least one non-residential place within walking distance for each category. This measure is denoted by *W_c_*and obviously, it is a discrete value and ranges between zero and four by definition. Second, we added up the total length of each walking trip with at most one trip from each category (if more than one non-residential place is within walking distance for one category, select the one with the shortest distance), and this measure is denoted by *W_d_*. Eventually, we varied the contexts to explore the corresponding results of the mean value of *W_c_* and *W_d_* across all households.

The first series of scenarios we explored do not exhibit spatial variation (i.e., the land use pattern was the same across the whole city) with a combination of the following dimensions: (1) the land use density was explored by various size of households in the city with a range of 1, 2, …, 20 units. Each unit represents 3,200 households; (2) we varied the ratio between the total number of households to the total number of non-residential places in each category with values of 25, 50, 100, 200, and 400; (3) the land use mix was explored by “zoning policy” implemented at five levels. As shown by Figure 1, at level one, the whole city was evenly divided to be four zones (2×2), and each category was assigned randomly to each zone (denoted by various colors), then all non-residential places belonged to a certain category were randomly distributed within that zone; at level two, each zone at the first level itself was further evenly divided to be four zones using the same process as mentioned above; similarly, level three, four, and five repeated the same processes and “go deeper” to a finer spatial scale. Correspondingly, the length of one side of a single zone (with a square shape) was 1600m, 800m, 400m, 200m, and 100m from level one to level five; (4) We explored the variation of walking distance with values of 400m and 800m.

**Figure 1.**
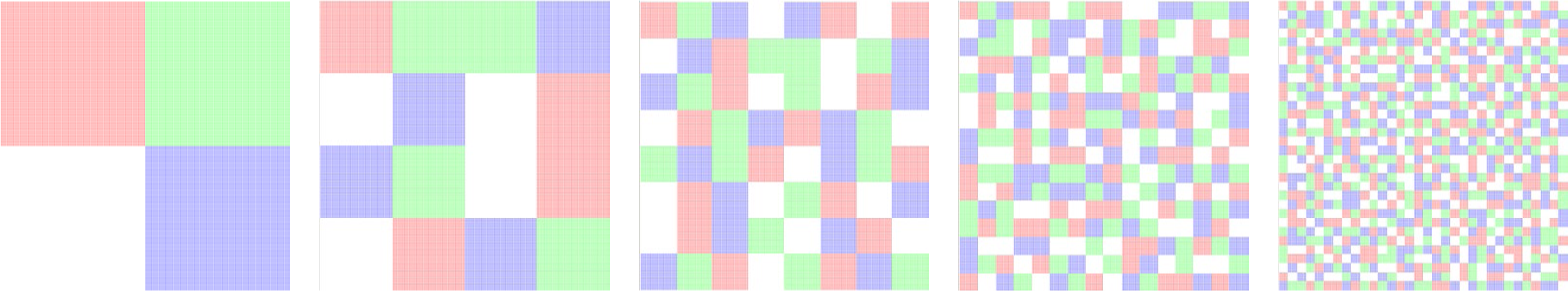
Examples of the schematic display of the land use mix within the city underlying zoning policy at five levels (from left to right are level 1 to level 5): each color indicates the zone for each category of non-residential places.

We designed a second series of scenarios to mimic that in reality the land use was not evenly distributed across the city. For example, non-residential places may be more likely to concentrate in the city center, non-residential places in the same category may be more likely to be clustered, and households may be located near or far from non-residential places. We explored four dimensions: (1) the land use density with 1-20 levels (see above); (2) spatial distribution of non-residential places with three options: random, one cluster in the city center, and four clusters (for each category) (see Figure 2); (3) spatial distribution of households with three options: random, one cluster in the city center, and four clusters (see Figure 2); (4) decay parameter for both the clustering of non-residential places and households, with four values of 0.001, 0.002, 0.005, and 0.01. We used a distance-decayed function *P*(*d*) = *e*^−β*d*^ to describe the relative probability of being located at a certain distance away from the cluster center. Distance decay functions [32–34] have been used to mathematically describe how the interaction between two locations declines as the distance between them increases. Specifically in this study, *d* is the distance from the cluster center, β is the decay parameter, and *P*(*d*) denotes the probability of this location to be occupied by a place. Table 1 shows the relative probability of being located for several selected distances using various values of the decay parameters. For example, with a decay parameter value of 0.002, a location 500 meters away from the cluster center is 0.368 times occupied by a place compared with the cluster center. In this series of scenarios, the ratio between households to non-residential places in each category was fixed to 100.

**Figure 2.**
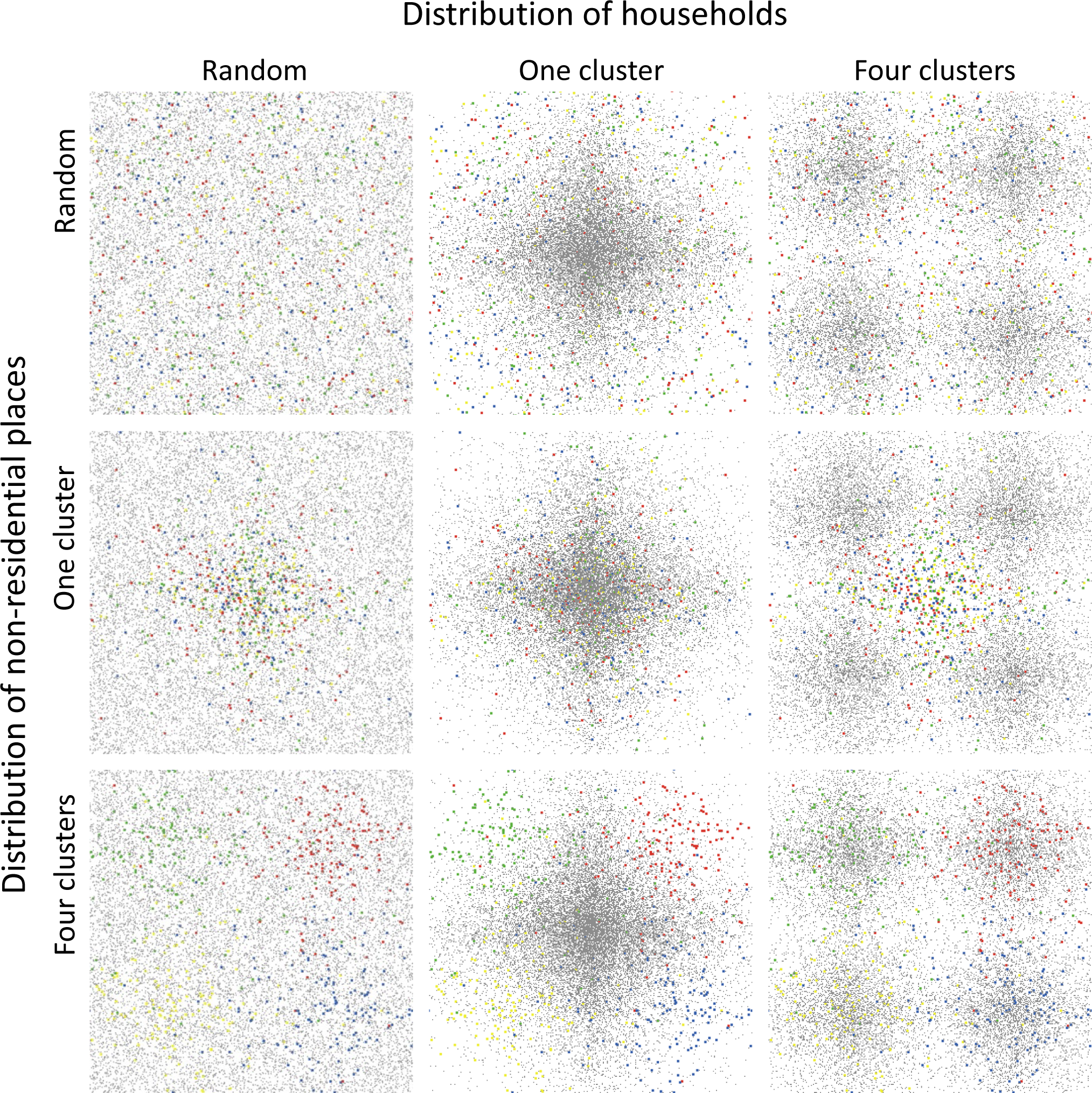
Examples of the schematic display of non-residential places and households within the city, and nine scenarios with the combination of (1) three options of the spatial distribution of households: random, one cluster in the city center, and four clusters (corresponding to the three columns from left to right); (3) three options of spatial distribution of non-residential places: random, one cluster in the city center, and four clusters (corresponding to the three rows from top to bottom). Households are denoted by gray dots and non-residential places are colorful squares with each color indicating each category. In this figure, all scenarios are with 0.002 for the decay parameter and 100 for the ratio between households to non-residential places.

**Table 1.** The relative probability of being located at several selected distances away from the cluster center by various values of the distance decay parameter.

| Distance from the cluster center | Distance decay parameter ($\beta$) | | | | |
| --- | --- | --- | --- | --- | --- |
|  | 0.0005 | 0.001 | 0.002 | 0.005 | 0.01 |
| 0 meter | 1 | 1 | 1 | 1 | 1 |
| 10 meters | 0.995 | 0.990 | 0.980 | 0.951 | 0.905 |
| 100 meters | 0.951 | 0.905 | 0.819 | 0.607 | 0.368 |
| 500 meters | 0.779 | 0.607 | 0.368 | 0.082 | 0.007 |
| 1000 meters | 0.607 | 0.368 | 0.135 | 0.007 | 0.000 |
| 2000 meters | 0.368 | 0.135 | 0.018 | 0.000 | 0.000 |

Among the second series of scenarios, we further examined the variation of W*_c_* and W*_d_* within the population. We selected scenarios with a fixed decay parameter of 0.002 and fixed ratio between households to non-residential places of 100, and examined the distribution of W*_c_* and W*_d_* within the population over the land use density, varying by both spatial distribution of non-residential places and spatial distribution of households.

## 3. Results

### 3.1. Scenario with evenly distributed land use

As shown in Figure 3, in all scenarios with evenly distributed land use, land use density was positively correlated with *W_c._* With the increase in density, *W_c_* increased fast at the beginning, then slowed down and nearly kept constant. With shorter walking distance (i.e., 400 meters compared with 800 meters) and a lower value of the ratio between household to non-residential, a higher threshold value of density is needed to reach the “nearly saturated” status (i.e., the *W_c_* stay constant or increase slowly with the increase of density). Higher land use mix level was related to a higher value of *W_c_*. The ratio between households to non-residential places did not significantly modify the relationship between land use mix level and *W_c_*. With a walking distance value of 400 meters, land use mix levels 4 and 5 had almost the same value of *W_c_*. While for walking distance value of 800 meters, the same value of *W_c_* resulted for land use mix levels 3, 4, and 5.

**Figure 3.**
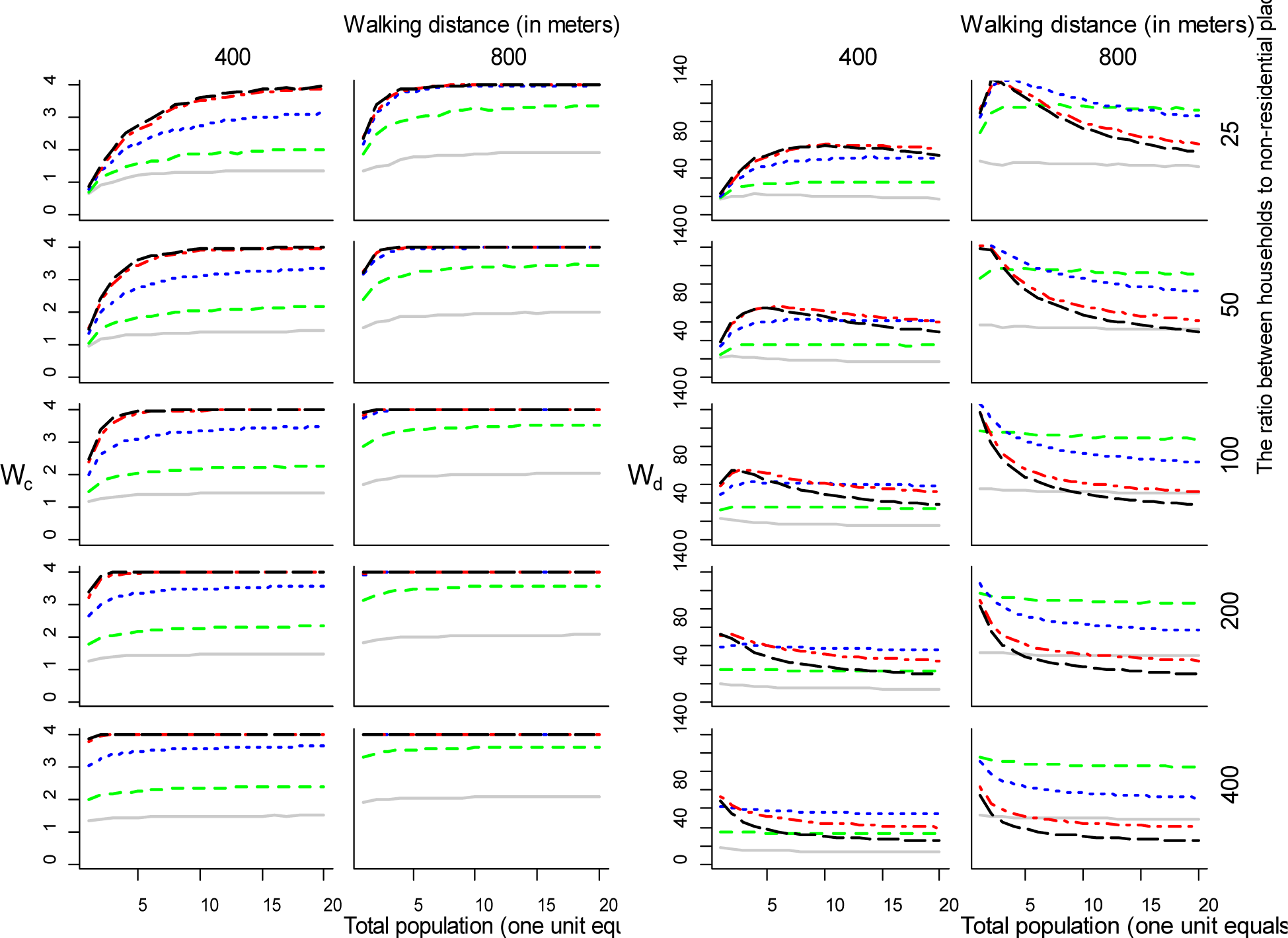
The mean value of *W_c_* (left panel) and *W_d_* (right panel) across all households over land use density (by X axis, each unit equals 3200 people), with various land use mix (by colors: gray, green, blue, red and black, from level 1 to level 5), walking distances (by columns), and the ratios between households to non-residential places in each category (by rows).

For *W_d_*, with a walking distance of 400 meters and 25, 50, and 100 of the ratio between households to non-residential places, with the increase of land use density, *W_d_*increased first, then came to be flat or decrease slowly. With a walking distance of 800 meters and 25 of the ratio between households to non-residential places, with the increase of land use density, *W_d_*increased first, then decreased significantly for a walking distance of 800 meters. Land use mix level was only positively correlated with *W_d_* when the walking distance was 400 meters and 25 or 50 of the ratio between households to non-residential places. When the walking distance was 400 meters and the ratio between households to non-residential places was 100, 200, or 400, land use mix level three resulted in the highest value of *W_d._*When the walking distance was 800 meters, for most values of ratios, it was land use mix level two resulted in the highest *W_d,_* and levels one and five resulted in the lowest *W_d_*.

### 3.2. Scenarios with spatially-patterned land use

As shown in Figure 4, when the decay parameter was 0.001, all nine curves were very similar in terms of both *W_c_* and *W_d_*. That is, with a very small value of decay parameter, walking was almost not influenced by both the spatial distribution of non-residential places and households. As shown in Table 1, with a smaller value of the decay parameter, the difference in the probability of being located at various distances away from the cluster center was smaller. Then regardless of the options for spatial distribution of non-residential places and households, the real spatial distribution of all places was similar to being randomly distributed across the city.

**Figure 4.**
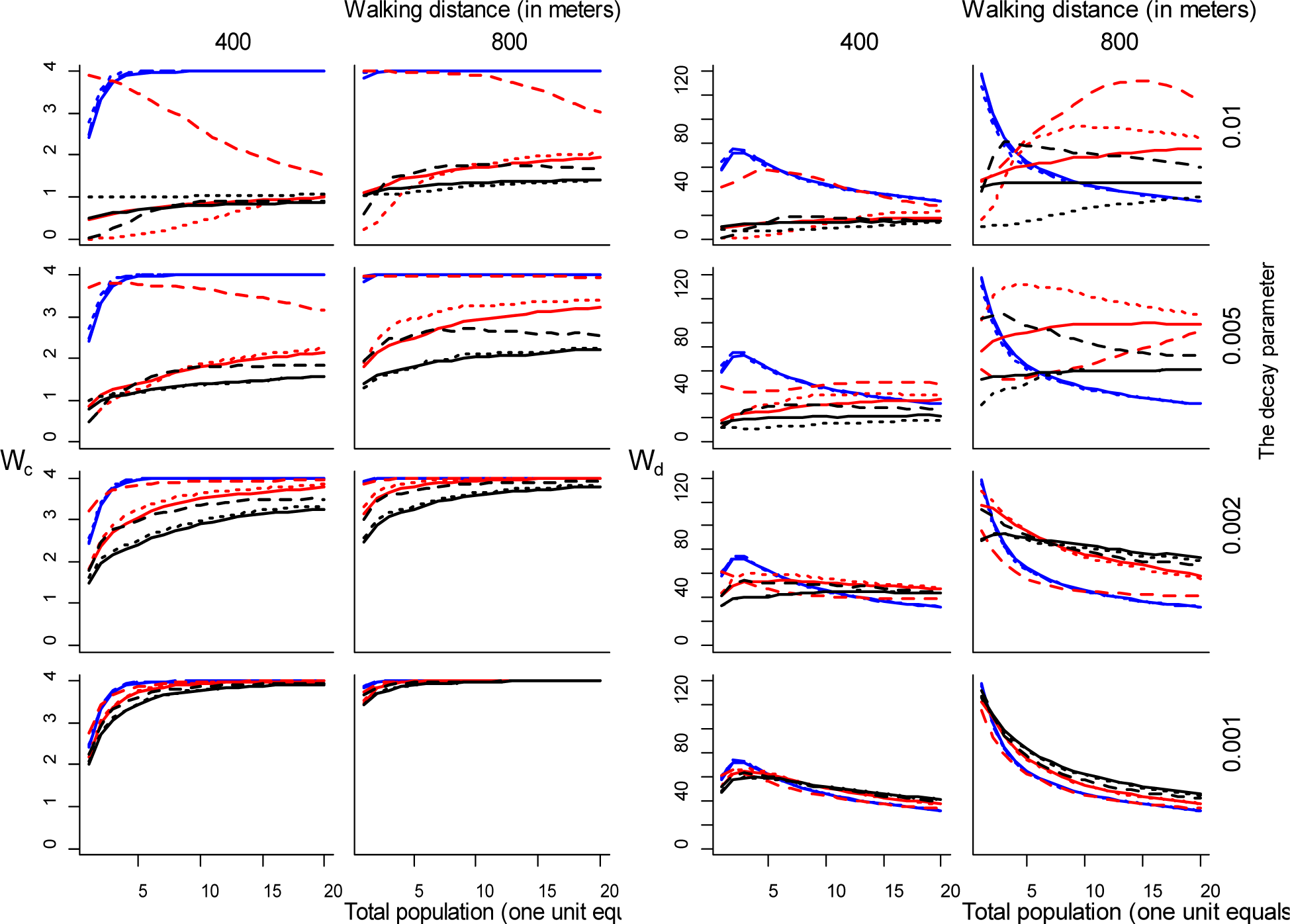
The mean value of *W_c_* (left panel) and *W_d_* (right panel) across all households over land use density (by X axis, each unit equals 3200 people), with various spatial distributions of non-residential places (by color, blue for random, red for one cluster in the city center, and black for four clusters), the spatial distribution of households (by curve style: solid line for random, dashed line for one cluster in the city center, and dotted line for four clusters), walking distances (by columns), and decay parameter for both the clustering of non-residential places and households (by rows).

As shown in Figure 5, all blue curves were similar regardless of the line styles. That is, when the spatial distribution of non-residential places was set to be random both *W_c_* and *W_d_* were almost not influenced by the spatial distribution of households.

**Figure 5.**
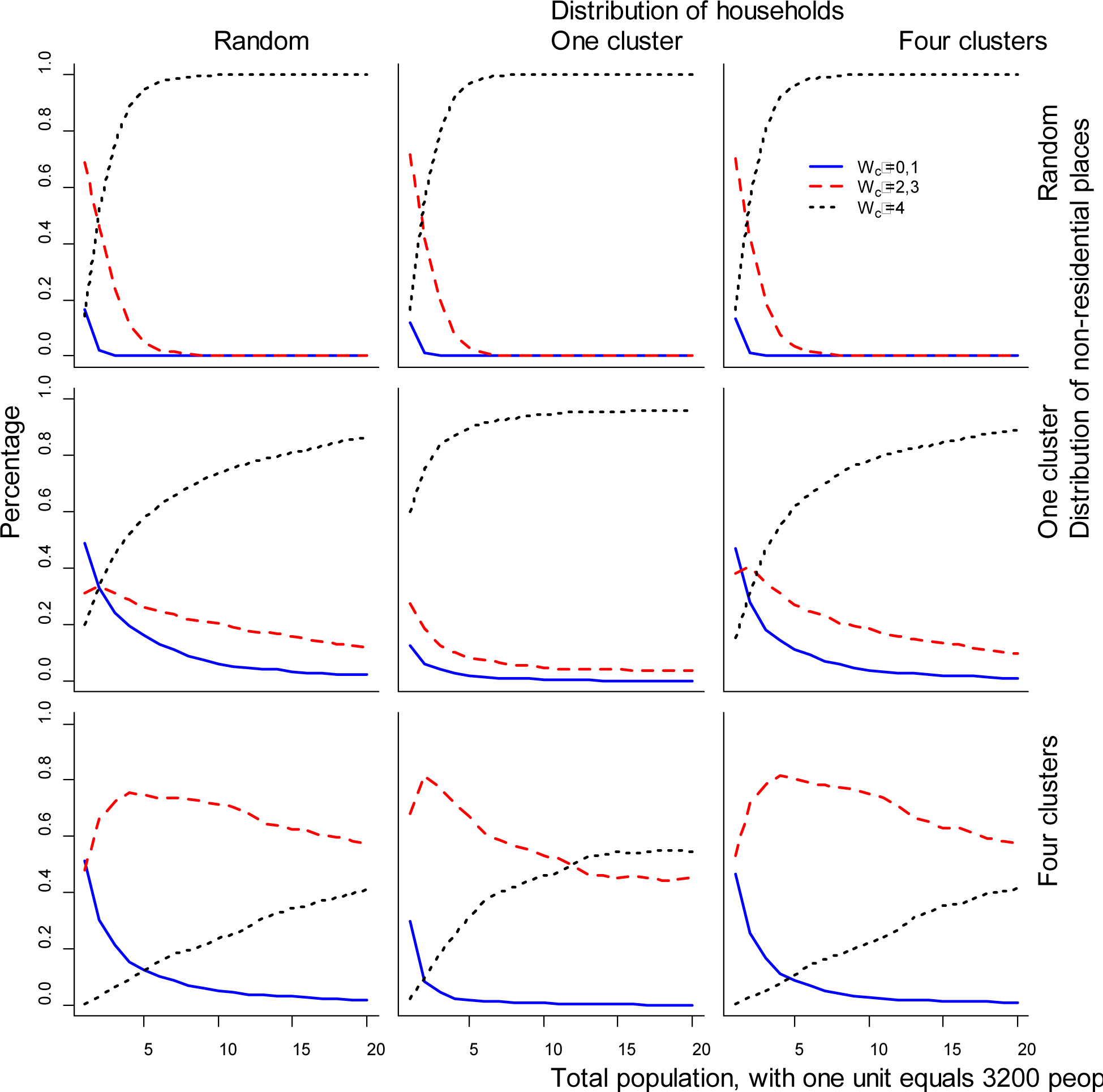
The distribution of *W_c_* (indicated by colors) within the population over the land use density (by X axis, each unit equals 3200 people), with various spatial distribution of non-residential places (by rows), spatial distribution of households (by columns). In this figure, all scenarios are with 0.002 for the decay parameter and 100 for the ratio between households to non-residential places.

For *W_c_*, in almost all scenarios, blue curves were higher than red curves, and red curves were higher than black curves. That is, for non-residential places, a randomly distributed pattern had with higher value of *W_c_* than one cluster, and one cluster was higher than four clusters. Further, within the groups of red and black, the dashed curves were higher than the other two curves. That is, for both one cluster or four clusters of the non-residential places, the one cluster of households in the city center had with highest *W_c_*. Almost all curves increased or increased first and then kept constant with the increase of the land use density, with a few exceptions: a red dashed curve in the highest value of decay parameter for both 400 meters and 800 meters of walking distance, the value of *W_c_* decreased with the increase of land use density. That is, with a fixed ratio of households to non-residential places, a higher value of land use density means a higher absolute number of non-residential places, and with a higher value of decay parameters, all these non-residential places are likely to be located in the city center, when the locations in the city center are nearly completely occupied by non-residential places. Then regardless of the one cluster pattern for the households, they have to be located further away from the center, forming a belt shape around the city center. Thus, higher land use density means a larger area of “core” consisting of non-residential places, and the households are “pushed” further away from the city center.

For W*_d_*, with 400 meters of walking distance, almost all curves increased first then began to decrease or kept constant over the increase of land use density. With 800 meters of walking distance, most curves kept decreasing, especially the blue curves, which decreased abruptly at the beginning and then slowed down gradually. The exceptions were red curves with higher values of the distance decay parameter. For most scenarios, with a lower density of land use, blue curves, that is, random distribution of non-residential places resulted in the highest value of W*_d_*; with a higher density of land use, red or black curves, that is, one or four clusters pattern resulted in a higher value of W*_d_*.

In Figures 5 and 6, the nine scenarios could be categorized by the distribution of non-residential places because the difference between categories is more significant than the difference within each category. In the following text, we presented the result by the distribution of non-residential places and switched between Figures 5 and 6.

**Figure 6.**
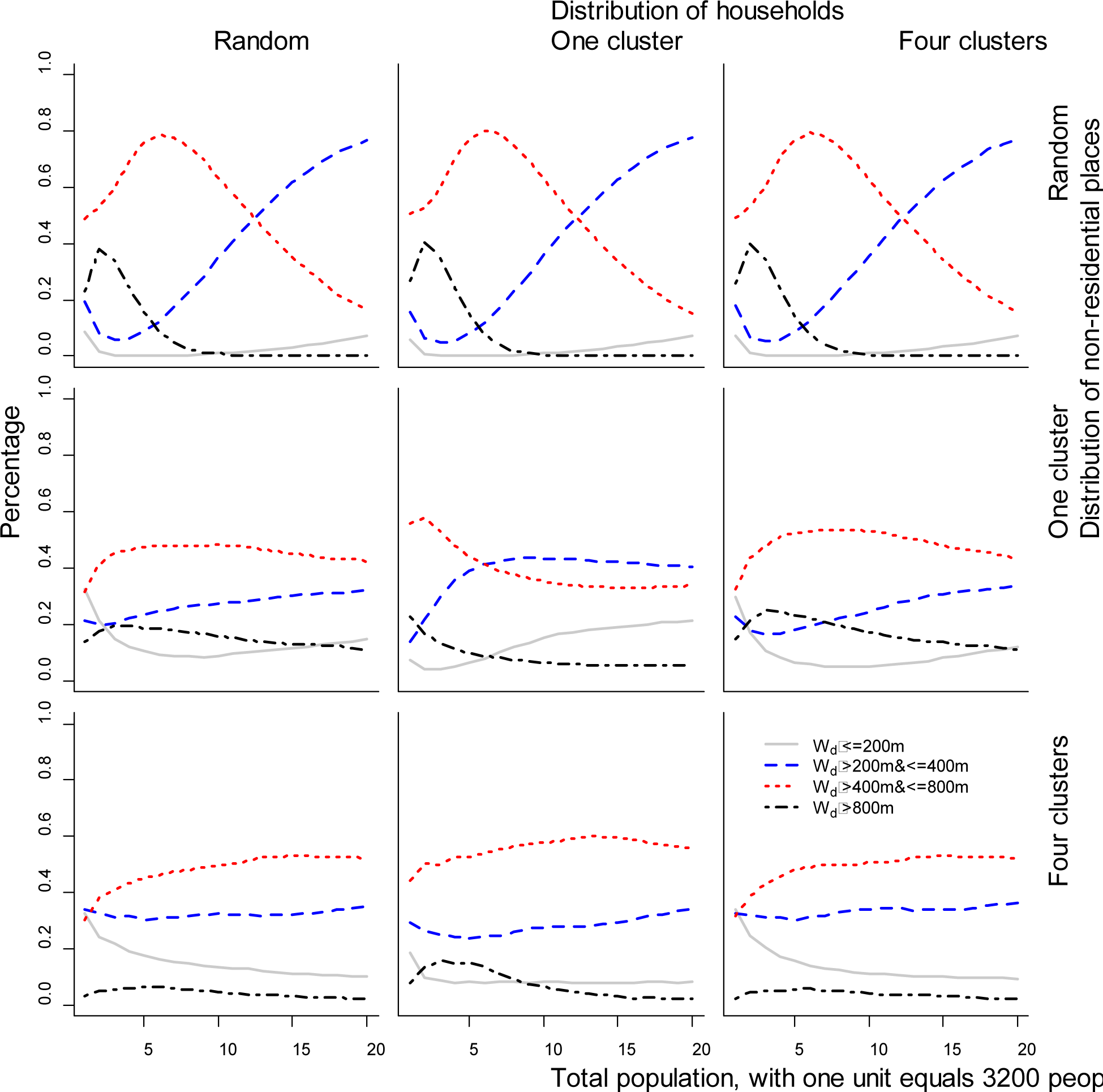
The distribution of *W_d_* (indicated by colors) within the population over the land use density (by X axis, each unit equals 3200 people), with various spatial distribution of non-residential places (by rows), spatial distribution of households (by columns). In this figure, all scenarios are with 0.002 for the decay parameter and 100 for the ratio between households to non-residential places.

With a random distribution of non-residential places and regardless of the distribution of households, the percentage of people with a maximum value of W*_c_* increased abruptly with the increase of land use density, until every person had at least one place in each category within walking distance. However, land use density was not positively correlated with W*_d_*. The percentage of people with W*_d_* more than 800 meters peaked at a very low land use density (about 2 units), and afterwards, it kept decreasing to zero. Also, the percentage of people with W*_d_* less than 800 meters but more than 400 meters peaked at lower land use density (about 6 units) and afterward kept decreasing as well. Over about 11 units of land use density, the majority of people were with W*_d_* less than 400 meters.

With one cluster distribution of non-residential places, the percentage of people with a maximum value of W*_c_* increased less abruptly than the random distribution of non-residential places, especially when the distribution of households was random or four clusters. Even at the highest level of land use density, still, not every person reaches the maximum value of W*_c_*. Generally, W*_d_* was less influenced by the land use density compared with random distribution, regardless of the pattern of households. With an increase in land use density, the percentage of people with W*_d_* more than 400 meters decreased slowly or kept constant, while the percentage of people with W*_d_* less than 400 meters increased slowly or kept constant.

With four cluster distributions of non-residential places and regardless of the distribution of households, a very small percentage of people with W*_d_* more than 800 meters, and people with W*_d_* between 200 and 800 meters increased gradually and people with W*_d_* less than 200 meters decreased.

## 4. Discussion

Using a very simple computer model, our study reveals how the land use density and land use mix may influence walking, at the most theoretical and fundamental level. As indicated, only at a very low level, increased land use density may increase *W_c_*. Over a threshold, increased land use density does not affect *W_c_*, and may decrease *W_d_* instead. The level of land use mix determines the full potential of *W_c,_* and *_a_* higher level of land use mix is associated with a higher value of *W_c._* However, a higher level of land use mix level is not associated with a higher value of *W_d_*. The relationship between walking and land use density and land use mix is significantly modified by factors such as walking distance and spatial pattern of land use within the city.

Our study confirms the non-linear relationship between land use density and walking and the existence of threshold value at the lower end of land use density which is contrary to some previous which found the threshold effect at the higher end of land use density. That could be explained by the clustering effect of higher land use density with other built environmental characteristics such as public transit and the synergistic effect between walking behavior and seeing others’ walking. Although higher land use density may increase walking and also decrease vehicle travel and fuel use [35], it should be noted that higher density may also increase social stress [27].

A higher level of land use mix means a higher probability of places with various categories near the household. However, if the level is too high, that means these non-residential places may be located too close to the household, so the walking trips are likely to become shorter. In this study, we used a special “zoning policy” to describe the diversity of land use mix at different levels. The change between levels is discrete instead of continuous. Measurement of land use mix is more complicated compared with land use density. As reviewed [11], measures of land use mix could be categorized as accessibility (the degree to which mixed-land activities are easy to reach by residents), intensity (volume or magnitude of mixed-land uses present in an area), and pattern (degree of evenness of various land-use types in an area). It is unclear which measures yield the strongest associations with the built environment and physical activity, and most measures do not correlate with all three dimensions. For example, the land use mix level in our study deals with accessibility exclusively, and intensity and pattern are not considered.

As our study indicated, the relationship between walking and land use density and land use mix is significantly modified by other factors, this may account for the mixed results as we reviewed in the introduction section. For example, we design a series of scenarios to represent the various spatial patterns of households and non-residential places. Because in real cases, in distribution of households and places are not evenly or randomly distributed. This partly reflects the mismatch of existing research on different spatial scales: variables of land use density or land use mix aggregated at a higher spatial scale do not represent effectively variation of the built environment at a lower spatial scale. Further, in this study, we used two values of walking distance as a reprehensive of the variation of walking among groups of people, while in reality, the variation of walking distance among the population is more complicated [34].

Even at a purely theoretical level, our model is very simple. Some assumptions the model is based on are rather arbitrary. First, we simplify the complicated land use into four categories with the same total size. Generally, land use categories include residential, commercial, institutional, industrial, recreational, and agricultural land uses. Although it is found that each category of non-residential place located within walking distance could result in five minutes of additional walking per week [23], it should be noted that not all categories of land uses contribute to walking for most people, and each category may vary for its importance to a certain group. As reviewed [4], including the land use category which has little or even a negative effect on people’s transport-related physical activity may obscure the measure’s relationship with physical activity outcomes. Indeed, the influence of land use on people’s physical activity not only varies by the land use category but also varies by people’s characteristics. Second, we assume that the distance is the only determinant of people’s travel mode choice, and further, it is simplified to be a dichotomous relationship while in reality people‘s tendency to walk decays over the trip’s length, and the decay function varies by trip purpose and subgroups [34]. In a larger picture, the relationship between the built environment and people’s active travel should also consider medium-term choices such as vehicle ownership and long-term choices such as residential and work location choices. Third, to simplify the model, some key characteristics of the built environment are excluded including street network, transportation system, and urban design.

Using a simple computer simulation, our research contributes to the fundamental knowledge about the relationship between the built environment and people’s physical activity. As reviewed [1, 11, 36], it is established that the built environment influences people’s physical activity in some important ways, although the causal relationship between the built environment and physical activity has not been established and we are not able to identify the strength of a specific feature. This is complicated because multiple factors at multiple levels could contribute to this relationship and the conclusions from the most existing practical studies may apply only to a specific population in a specific context. In this regard, a simple computer model will be supplemental because it provides a method to disentangle the complex relationship by separating and removing confounding factors and focusing on the interesting factors. In this regard, we may need more advanced simulations to explore policy interventions, because effective policies vary by different purposes of physical activity, by different population groups, and by different contexts.

## Data Availability

All data produced in the present study are available upon reasonable request to the authors

